# How should menorrhagia be managed in people with bleeding disorders: A systematic review of the literature and thematic synthesis

**DOI:** 10.1101/2025.03.22.25324427

**Authors:** Rameen Masood, Vidiya Dev, Millie Gee, Katie Finch, Daniel M. Fletcher, Olufikayo Bamidele, Jo Traunter, David Allsup, Barbara-ann Guinn

## Abstract

**Objectives:** To determine how menorrhagia is managed in people with bleeding disorders.

**Design:** A systematic review and thematic synthesis.

**Data sources:** PubMed, Medline, Scopus, Cochrane library, google scholar and CINAHL complete (via EBSCO).

**Methods:** Searches were conducted on articles published from 1^st^ January 2000 until 6^th^ May 2024. Following deduplication, the titles and abstracts were screened for relevance. 244 primary studies were then assessed for eligibility based on inclusion and exclusion criteria. Studies were included if they were based on primary articles and focussed on people with inherited bleeding disorders and heavy menstrual bleeding. Included studies were appraised for risk of bias and quality assurance using the Newcastle Ottawa Scale, following which data was systematically coded to generate descriptive and analytical themes.

**Results:** We identified 16 eligible articles of which 13 were included in a thematic synthesis. These included prospective and retrospective clinical studies, cross-sectional studies and randomised control trials encompassing over 893 participants. Thematic synthesis identified hormonal treatments, such as the levonorgestrel-releasing intrauterine system (LNG-IUS), to be largely effective in the symptom management of HMB in IBD and associated with improved quality of patient life.

Treatment of HMB patients with LNG-IUS, followed by tranexamic acid (TA) or 1-deamino-8-d-arginine vasopressin (DDAVP), the trade name for desmopressin, commonly led to amenorrhea. Technological approaches to the management of HMB in IBD included the use of mobile technology to encourage treatment compliance. These management strategies led to an improvement in reported QoL by patients with IBD. This review had limitations including the exclusion of some articles that may have limited generalisability. The Medical Subject Heading (MeSH) terms used focussed on HMB as opposed to abnormal menstrual bleeding, potentially directing the identified recommendations for clinical practice. Based on the findings of this thematic review, the use of LNG-IUS as first line therapy for those with HMB, followed by the use of combination therapy such as TA and desmopressin, would be recommended. These measures should be adopted in both primary and secondary care settings. We identified the need to strengthen counselling and communication between specialists involved in the care of those with HMB and IBD, and the need to increase awareness of HMB in IBD through public and patient education.

**Data availability statement:** All data presented is secondary to published studies and available within the public domain.

**Registration:** PROSPERO registration number: CRD42023452533

**Key Messages:**

- **What is already known on this topic** – Heavy menstrual bleeding (HMB) is often a symptom of inherited bleeding disorders (IBD) in females and can have a significant impact on the quality of life of an individual.
- **What this study adds** *–* A systematic review and thematic analysis of the currently available literature allowed the identification of best practise management options for patients with IBD and HMB. A thematic synthesis was used to identify best practice for IBD patient treatment and management of HMB, which will improve patient quality of life.
- **How this study might affect research, practice or policy** – This study of existing literature and thematic synthesis has been used to provide recommendations to haematologists and gynaecologists to support evidence based best practise recommendations on how to treat patients with HMB consequent to IBDs.

## INTRODUCTION

Menorrhagia is excessive menstrual blood loss that can interfere with a physical, social, emotional, and material quality of life (QoL) (1). Menorrhagia has been quantified as blood loss of more than 80 ml or occurrence of bleeding over a duration of more than 7 days but it is widely recognised that a diagnosis of menorrhagia is highly subjective (2). The International Federation of Gynaecology and Obstetrics (FIGO) divided the causes of menorrhagia or Heavy Menstrual Bleeding (HMB) into structural and non-structural categories (3). FIGO noted the non-structural cause of HMB to be coagulopathy, notably, inherited bleeding disorders (IBD). It is known that 20 percent of adolescent females who present to primary care with HMB will have a bleeding disorder, yet most of these females will not undergo testing for an IBD (4).

IBDs affect 1 in 2000 people in the UK (4) with Von Willebrand’s Disease (VWD) being the most common. VWD has a prevalence of estimated 16.5 people per 100000 in the UK population (5). IBD can be symptomatic with HMB, while HMB has been described by many individuals with IBD as having the largest negative impact on their QoL, when considering all of their symptoms (6).

Although scientifically there is some awareness of the relationship between HMB and IBD, this does not appear to translate into clinical practice. The European Association for Haemophilia and Allied Disorders (EAHAD) state that the underdiagnosis of IBD in females is common and that women typically experience diagnostic delays of between 8 and 16 years (7). EAHAD outline ten principles that aim to improve health, wellbeing and QoL in females with IBD. These principles highlight the need for clear pathways from diagnosis to treatment (4).

With regards to managing HMB, The National Institute for Health and Care Excellence (NICE) offers guidance for those without an underlying pathology, such as IBDs, and to those with fibroids (1). In these patients NICE recommends the use of hormonal treatment, tranexamic Acid (TA), and non-steroidal anti-inflammatory (NSAID) medication to manage patients with fibroids less than 3cm and with suspected or diagnosed adenomyosis. For HMB with larger fibroids, NICE has more tailored management recommendations (1). There is an absence of NICE published guidance for the management of HMB in those with IBDs. Additionally, FIGO have not supported any published recommendations on how to manage HMB in IBD. As discussed, FIGO have only published systems to understand menstrual bleeding to aid clinicians in developing management plans for those with HMB (8).

A UK Haemophilia Centres Doctors’ Organisation published guidelines for the gynaecological management of women with IBD in 2022, intended for haematologists and gynaecologists involved in the care of those with HMB and IBD. They recommend the use of TA in preference to desmopressin to prevent and treat HMB in certain IBDs. Regarding hormonal treatment, the use of the LNG-IUS is recommended as an effective first line therapy for HMB in IBD for those who do not wish to conceive (9).

Not all patients with an IBD require active management with pro-haemostatic therapies, with such interventions often only required in response to trauma or surgical procedures. The British Society for Haematology recognise, that in some people with VWD, prophylactic therapy is needed to reduce the frequency of bleeding episodes (10). However, whilst prophylaxis for bleeding episodes does appear useful in decreasing bleeding events there appears to be a knowledge gap regarding the efficacy of this therapy on HMB (11).

Therefore, the aim of this study was to use evidence based clinical findings to define the best management of HMB in those with IBD. To achieve this a systematic review of the literature was performed.

### Objectives

- To determine the clinical experience(s) of people with bleeding disorders and HMB a stakeholder survey was utilised.
- To support these findings from a broader group of patients a systematic review and qualitative evidence synthesis of patient’s experiences were used to build a rationale for why and where a primary study might be needed.

## METHODS

### Public and Patient Involvement

This study was initiated by a patient with an IBD and experience of HMB. Their priorities, experience and preferences informed our research question. They were engaged throughout the study and helped inform the inclusion and exclusion criteria. They were given time to engage with the data extraction and read the article as it developed into its final format.

### Systematic Review

This systematic review adheres to the Preferred Reporting Items for Systematic Reviews and Meta-Analyses (PRISMA) guidelines (12) including the development of a Protocol dataset Supplementary Information I and prospective registration with PROSPERO (CRD42023452533).

### Eligibility criteria

The research question was developed using the following PECO framework (13):-

- **P**articipants - adults with HMB and IBD.
- **E**xposure - treatment to manage HMB.
- **C**omparators - patients whose HMB was not managed
- **O**utcome - impact on QoL of the patient.

Studies assessed were not limited to those in the English language but included participants with experience of HMB and IBD; with no age, geographical location, publication date, or setting restrictions (Table 1). Papers were excluded if HMB was absent and/or bleeding disorders were not diagnosed or were from a non-genetic aetiology. In addition, reviews and studies of less than ten participants/case reports were excluded.

**Table 1.** Exclusion/inclusion criteria.

| Inclusion | Exclusion |
| --- | --- |
| Primary literature sources: with reviews kept in until screening is completed for the purpose of backward snowballing | Case studies, reviews |
| Human studies | Studies on cell lines, animal models of inherited bleeding disorders or heavy menstrual bleeding |
| Patients diagnosed with heavy menstrual bleeding and a bleeding disorders | Disease other than heavy menstrual bleeding<br>Disease other than inherited bleeding disorders |
| All ages |  |
| All geographical locations |  |
| All publication dates |  |
| All languages |  |

Review articles were only removed once the citations in all selected manuscripts had been screened against the inclusion/exclusion criteria. This ‘backward snowballing’ step helped ensure that relevant literature was successfully found as part of a systematic review.

### Information sources

We identified articles from six online databases (PubMed, Medline, Scopus, Cochrane, google scholar and CINAHL) from 1^st^ January 2000 until 6^th^ May 2024.

### Search Strategy

The development of the search strategy dataset was based on the index terms found in three to six sentinel articles following an initial PubMed screen of the literature. Published manuscripts focusing on HMB and IBD were identified using MeSH search terms and variations thereof as follows: ((exp Blood Coagulation Disorders) OR (Clotting disorder*)) AND ((exp Menorrhagia) OR (heavy period) OR (Menorrhagia) OR (Heavy menstrual period)). These searches were repeated across all six databases.

### Selection process

Article selection undertaken by two independent pairs of screeners; Group 1: R.M. and V.D. up to September 2023, and Group 2: M.G. and K.F. from September 2023 to May 2024. Both pairs of screeners discussed any queries regarding differences in selected publications. Excel was used as a repository for articles and following deduplication, the articles were screened by title, abstract and full text against the inclusion and exclusion criteria (**Table 1**).

### Risk of bias assessment

The Newcastle Ottawa Scale (NOS) for Assessing the Quality of Nonrandomized Studies in Meta-Analysis was used to perform quality assurance assessment of the selected studies (14). The studies were assessed based on selection, comparability and outcome, and were ranked from zero to four stars. Zero stars signified a lack of information required, and four stars signify perfect match to the criteria, and with no indication of what could be improved upon.

### Data extraction

For standardisation, a data extraction form was piloted on Excel using several selected studies with input from all reviewers. This included fields for study methodology, interventions/managements being reviewed, sample size, qualitative experience. The pairs of screeners performed data extraction on their selected and agreed articles.

### Thematic Synthesis

Based on previous guidance (15), we performed the thematic synthesis of data in three stages. Data from each paper was coded and reviewed to identify key concepts, using text directly from the selected articles. Concepts included the management options and recommendations regarding best practice in the management of HMB and IBD, as presented by each paper. Similar concepts within and between papers were grouped to create descriptive themes. Reviewers weighted themes according to the frequency in which they appeared across the articles. The greater number of times a theme was identified, the more weight was given to it as a theme. Comparisons between themes were made and these were subjectively interpreted in accordance with our research intentions to synthesise analytical themes. This was conducted collaboratively by three reviewers (V.D. R.M. and M.G.) using the excel data extraction form.

## RESULTS

### Screened studies selected for systematic review

The search amassed 11568 papers, of which 5671 were duplicates (Figure 1). We used a prespecified inclusion/exclusion criteria (Table 1) to screen articles on title and abstract resulting in 244 articles being selected. At the stage of abstract reading our search excluded books, systematic reviews, meta-analyses and conference papers. The articles were then screened by full text, with 16 meeting the eligibility criteria to be included in further analysis (Supplementary Information II). When it came to full text analysis, papers with only abstracts and those that didn’t meet the inclusion/exclusion criteria were removed.

**Figure 1.**
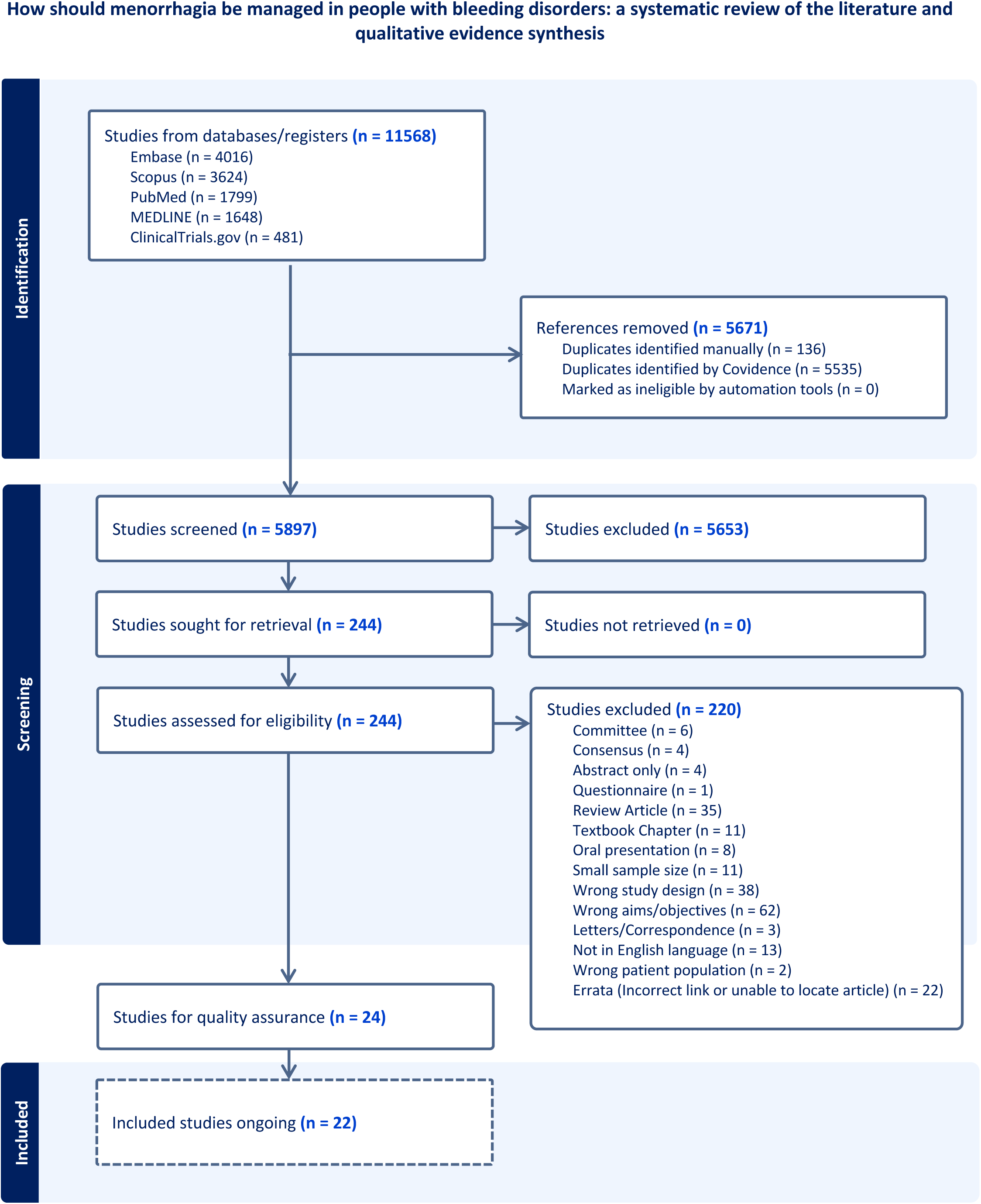
Prisma-P flow diagram. Flowchart of the screening process for articles which had considered the management of people with heavy menstrual bleeding and bleeding disorders.

Reviews remained in the systematic review until it was completed, these were then used for a backward snowballing step (16) (Supplementary Information III) and we found 35 articles that had been excluded during our original searches which were then fully assessed. Any disparities in chosen articles were discussed between the reviewers and a consensus achieved. However, when disagreements remained, they were resolved by a third reviewer (B.G.). Articles that could not be accessed were sought from the corresponding author and, where different, the senior author.

### Quality assurance

The full-text versions of 16 studies on HMB and IBD were assessed for eligibility and quality using a modified NOS template (Table 2). Three of them were excluded considering the risk of bias and applicability (Table 3) and all others met the requirements for data extraction (Supplementary Information IV). This identified 13 studies that met the quality assurcancer criteria for data extraction.

**Table 2.** Selection, Comparability and Outcome assessment applied to the selected studies. NOS guidelines for cohort studies were followed. Studies with zero stars were subsequently excluded from further meta-analysis.

| Reference | Selection ( /4) | Comparability ( /2) | Outcomes/Exposure ( /3) | Rating |
| --- | --- | --- | --- | --- |
| Campos et al., 2020 | *** | * | *** | Good |
| Chi et al., 2011 | ** | * | ** | Fair |
| deWee et al., 2011 | ** | ** | - | Poor |
| Dietrich et al., 2017 | ** | * | ** | Fair |
| Dowlut-McElroy et al., 2015 | *** | * | * | Poor |
| Govorov et al., 2021 | *** | * | ** | Good |
| Greenmyer et al., 2021 | ** | * | - | Poor |
| Huguelet et al., 2022 | *** | * | ** | Good |
| Huq et al., 2011 | *** | ** | ** | Good |
| Kadir et al., 2002 | *** | * | ** | Good |
| Leissinger et al., 2001 | *** | ** | ** | Good |
| Pennesi, 2020 | *** | ** | ** | Good |
| Ragni et al., 2023 | *** | ** | ** | Good |

**Thresholds for converting the Newcastle-Ottawa scales to AHRQ standards (good, fair, and poor):**
*Good quality:* 3 or 4 stars in selection domain AND 1 or 2 stars in comparability domain AND 2 or 3 stars in outcome/exposure domain
*Fair quality:* 2 stars in selection domain AND 1 or 2 stars in comparability domain AND 2 or 3 stars in outcome/exposure domain *Poor quality:* 0 or 1 star in selection domain OR 0 stars in comparability domain OR 0 or 1 stars in outcome/exposure domain

**Table 3.** Articles removed during QA and RoB analysis.

| Reference | Reason for Exclusion – Poor rating |
| --- | --- |
| deWee et al., 2011 | This was a cross-sectional study and bleeding severity was measured using a given score |
| Dowlut-McElroy et al., 2015 | No real evidence of follow-up with the cohort |
| Greenmyer et al., 2021 | Could possibly touch upon this new type of management which can be considered for treating heavy menstrual bleeding in patients with bleeding disorders |

### Data extraction

In the 13 studies, a total of 1069 participants were included. The largest study involving 423 participants in Netherlands, and the smallest study had 11 participants and was conducted in the USA (19, 28). Seven studies were undertaken in the USA (17–23), three in the UK (24–26), one study in Brazil (27), one in the Netherlands (28) and one in Sweden (29). There were no studies that only examined qualitative data, however six studies performed quantitative analysis only (18–20, 22, 27, 28)(Supplementary Information IV) and seven studies conducted both qualitative and quantitative analyses (17, 23–26, 29). The qualitative analysis focused on discussing lived experiences of patients and the quantitative analyses primarily determined scores for the Pictoral Blood Assessment Chart (PBAC) (23–27, 29), blood tests (25, 27), QoL (21, 23–29) and/or other measures (18–20, 22) (Supplementary Information VA).

### Thematic Synthesis

Patients analysed in these clinical settings expressed a range of emotions concerning the management of menorrhagia in the context of their bleeding disorders. Four key analytic themes were identified reflecting patients’ experiences with menorrhagia and bleeding disorders during diagnosis, treatment and symptom management (Supplementary Information VB & VC).

#### 1. Restoring Control: Tackling HMB

For women with IBD, HMB is more than just a clinical issue and in many cases served as a persistent disruption to daily life. The advent of the levonorgestrel-releasing intrauterine system (LNG-IUS) has provided many women with renewed hope enabling them to regulate their cycles, significantly mitigating blood loss and enhancing overall quality of life. Five studies (19, 24, 27–29) within this systematic review have affirmed that LNG-IUS is highly effective in managing HMB.

Campos et al (27) reported that ‘The use of LNG 52-mg IUS was effective in reducing menstrual bleeding in the women with IBD by 3- months from baseline’ and further mentioned ‘The median PBAC score was higher before LNG 52-mg IUS placement than at 3, 6, and 12 months after placement (p<0.001), demonstrating its efficacy’.

Chi et al (24) addressed the practicalities of introducing the LNG-IUS as a management option for individuals with HMB and IBDs, stating that the LNG-IUS ‘can be easily inserted in a clinic setting and is well accepted by the women. It should be offered to women with IBDs who also require contraception and certainly prior to surgical options’.

#### 2. Non-hormonal treatment and other therapies

Non-hormonal therapies provided an important alternative for managing menorrhagia in patients with IBD, particularly for those who cannot tolerate or prefer to avoid hormonal treatments. Ten studies (18, 19, 21–28) explored a wide range of non-hormonal and alternative treatment strategies, including desmopressin, TA and iron supplementation and offering insights into their mechanisms, efficacy, and role in personalised patient care.

Combination therapy is an alternative that can be considered for managing HMB, benefitting patients by integrating both hormonal and non-hormonal agents to achieve efficient symptom control. Dowlut-McElroy et al (18) recommended ‘that strong consideration be given to combination non-hormonal and hormonal modalities for the treatment of HMB in girls with bleeding disorders’.

For women who do not wish to preserve their fertility, endometrial ablation could be considered. Huq et al (25) concluded that ‘Endometrial ablation appears to be a safe and effective long-term treatment for HMB in women with IDBs. It significantly decreases menstrual blood loss and improves QOL’.

DDAVP was explored as an option to manage and prevent HMB in IBD. Leissinger et al (21) suggested that ‘when used for the treatment of menorrhagia, the efficacy of high-dose DDAVP intranasal spray (1.5 mg mL−1) was rated as ‘excellent’ after 655 (92%) of 721 daily uses’. However, Kadir et al (26) also explored the use of the DDAVP nasal spray, but found no statistically significant improvement in the management of HMB in IBD.

Recombinant von Willebrand Factor (RVWF) is another non-hormonal treatment option worthy of consideration. Ragni et al (23) compared the effectiveness of RVWF and TA for the management of HMB and found that ‘rVWF is inferior to TA in reducing HMB in subjects with mild or moderate VWD, with neither treatment showed a clinically relevant effect’.

#### 3. Reclaiming Quality of Life: How to Shift from Struggle to Strength

HMB not only causes physical distress but also profoundly impacts various facets of a woman’s life, including career choices, mental health and social participation. The studies analysed in this systematic review demonstrate that LNG-IUS significantly improves quality of life (QoL), with notable improvements in physical functioning, emotional well-being, and pain reduction. In addition, patients with menorrhagia are at an increased risk of developing iron deficiency anaemia, a chronic condition that can profoundly impact their quality of life. The resulting fatigue and, in some cases, cognitive impairment can further exacerbate daily challenges, limiting their ability to perform routine activities and affecting overall well-being.

Therefore, the ability to resume activities of daily living (ADLs) without constant worry about heavy bleeding represents a transformative change for many patients.

Eight studies (22–29) utilised the Rand 36-item short form survey (SF-36) which scores eight health concepts: physical function, bodily pain, role limitations due to physical health problems as well as personal or emotional problems, emotional well-being, social functioning, energy/fatigue and general health perceptions as well perceived change in health. Quality of life was found to be improved through the insertion of LNG-IUS as follows:- ‘There was an improvement in all eight parameters of quality of life (p<0.001). The mean haemoglobin, ferritin, and serum iron levels were also higher at 12 months than before LNG 52-mg IUS placement’ (27). These results indicate that patients with HMB may commonly have iron deficiency without anaemia, as proposed by others (30). Huq et al (25) highlighted the improvement in QoL after endometrial ablation: “The overall scores for all categories included in the QOL improved significantly after treatment (P<0.01). Prior to ablation, 67% rated their general health as poor compared with 0% at follow-up”.

Pennesi et al (22) stated that ‘Young women with HMB miss more days of school and have increased disruption of hobbies and activities and decreased sports participation because of menses, compared with their peers with normal menses’. This typifies how HMB negatively impacts the lives of individuals with IBD and that managing HMB in those with IBD contributes to an improvement in their QoL.

#### 4. Rethinking Menstrual Health: A Call for Better Awareness and Research

Women’s health, particularly in the field of menstrual conditions and IBD, remains an underfunded and under-researched area. Seven studies (17, 18, 22, 23, 25, 28, 29) have highlighted the importance of increased education and awareness. A range of management options should be considered and the most efficacious chosen based on individual symptoms and underlying pathology.

One of the strategies being assessed is the use an iPod Touch (Apple Inc, Cupertino, CA) device (ITD), to monitor treatment compliance levels. ITD is predicted to help ‘improve educational access through engaging technology’ (17). ITD can be introduced and disseminated to various centres; and used as a useful tool for ‘managing adolescents with HMB and BDs by virtue of its inherent adaptability, minimal need for training, low cost, and potential to improve patient compliance with treatment regimens’ (17).

Good communication between clinicians involved in the care of patients with HMB and IBD was highlighted, as “close liaison with the Haemophilia team is essential to determine the risk of bleeding and the need for haemostatic prophylaxis” (25). The article described the need for management to be a discussion between haematology and gynaecology or the general practitioner to provide best care. It was proposed that communication needs could also be met by ‘the creation of a national registry that standardizes the collection of menstrual data and treatment plans would further delineate menstrual patterns and effective treatment of HMB in girls with bleeding disorders’ (18).

Counselling patients of their treatment options, was a common recommendation, noted particularly in three articles (18, 22, 29). A significant number of individuals with HMB and IBD appear not to be involved in the discussion surrounding their own management options, when a plan was being developed. Of note, ‘Only 28% of the cohort had notations indicating that a treatment plan for HMB had been discussed with their provider prior to menarche (P = .0001)’ (18).

## DISCUSSION

To address our research question, we performed a systematic literature review of the management of HMB in patients with IBD that identified thirteen full text articles as meeting our criteria. We found four key themes amongst these articles, highlighting various treatment modalities and their effectiveness.

LNG-IUS (19, 24, 27–29) was the most common treatment given to patients, and highlighted its importance as a long-term approach due to its dual benefits of reducing bleeding and potentially decreasing the requirement for repeated, more invasive interventions. Additionally, these articles (19, 24, 27–29) emphasised that LNG-IUS offered a better patient experience as it enabled greater autonomy and convenience due to its minimal requirement for maintenance. Patients appreciated that the IUS could remain in place for many years, particularly beneficial for patients who required the continuous management of HMB.

One additional point of interest relates to the expulsion of the IUD before the completion of the treatment period, highlighted in the literature. It is worth considering that this could potentially introduce bias into treatment outcome assessments, as early removal of the IUD may have resulted in incomplete treatment or inadequate follow-up data, impacting the overall effectiveness and comparison of treatments. Through analysing this treatment method, several recommendations can be implemented in clinical practice to ameliorate patient experiences and efficacy of treatment. It is advisable to consider LNG-IUS as a first-line treatment for HMB. Gynaecological guidelines need to be updated, and it is essential to critically consider whether long-term oral medications are leading to beneficial outcomes or whether it would be more advantageous to introduce LNG-IUS intervention at an earlier stage. In addition, future research should focus on developing personalised treatment protocols, optimising insertion techniques, and identifying patient subgroups who may benefit most from LNG-IUS therapy. In addition, it is crucial to improve access to this treatment option, especially for those women who have historically been dismissed or left with few viable solutions.

Leissinger et al. (21) examined the subjective patient experience with Vasopressin/Desmopressin nasal spray, focusing on patient-reported satisfaction rather than providing quantitative clinical outcomes. The absence of objective treatment outcomes makes it difficult to draw definitive conclusions regarding the clinical efficacy of these treatments. The efficacy of TA and desmopressin nasal spray seemed to be associated with positive outcomes for individuals with HMB and IBD (21, 26). TA, an antifibrinolytic, is widely used for its affordability and minimal side effects. One of the studies (23) compared rVWF and TA through a randomised crossover trial. The study found that while both treatments reduced PBAC scores and the frequency of flooding, rVWF achieved a slightly smaller reduction in HMB by absolute PBAC score compared to TA. Although neither treatment was able to normalise PBAC scores completely, both made significant improvements in symptom severity. Importantly, QoL and healthcare utilisation were comparable between the two groups. Given its cost-effectiveness and comparable efficacy, TA emerged as a more practical option for HMB management, particularly for patients looking for a balance between treatment impact and affordability. Endometrial ablation and octreotide treatment (19, 25) were also demonstrated to have varying degrees of success in addressing menorrhagia in VWD patients. These studies did not include direct comparisons with other therapies and therefore lacked the comparative data necessary for a broader analysis of their relative efficacy.

More innovative approaches to management and to monitor compliance were also considered, including the use of ITD to track and manage HMB in IBD (17). Participants utilised the ITD to record information about their IBD based on what they learned from IBD-related websites, as well as track their menstrual cycles and medication usage. This electronic data, coupled with charts of records, provided a closer insight into compliance with prescribed treatment regimes. According to the study, this self-monitoring approach guided and supported the patients to remain pro-active in managing their symptoms, promoting better adherence to treatment, and allowing adjustments based on real-time data. Nonetheless, the limitation with this study was that it did not report relevant treatment outcomes. And whilst the outcomes from this study are important, they do not directly address the effectiveness of specific treatments in managing menorrhagia, thus excluding the study from a comparative analysis of treatment efficacy.

Three studies (20, 24, 27), provided data on the proportion of patients achieving amenorrhoea following treatment. Campos et al. and Chi et al. (24, 27) did not quantify the amount of bleeding before and after treatment while Huguelet et al (20) provided an in-depth report on the rate of amenorrhoea among young adults and concluded that in individuals with high rates of amenorrhea, LNG-IUS is an effective treatment consideration. Pennesi et al. (22) failed to provide specific data, and the study was ultimately deemed unusable for inclusion in the meta-analysis. Furthermore, the study by Huguelet et al. (20) was paywalled, and despite efforts to access the data, we were unable to obtain the necessary information for review, which further limited the available evidence.

Currently, there are no readily available guidance regarding the optimal management of individuals with IBD that present with HMB. The NICE guidelines (1), which are widely used by gynaecologists for managing HMB, offer only two non-hormonal treatments namely TA and NSAIDs. Hormonal treatments include combined hormonal contraception and cyclical oral progestogens. Surgical options include second-generation endometrial ablation and hysterectomy. Moreover, guidance, presently, only supports the investigation of coagulation disorders in those who present with HMB, without a current diagnosis of an IBD.

One of the studies (18) compared the efficacy of hormonal treatment with non-hormonal treatment, elucidating that, while hormonal therapy was used as the initial treatment method for just over half the affected population, combination approaches were favoured for on-going treatment. This demonstrated that there is a need for more than one treatment strategy to address the severity of HMB in many cases, as discussed in our recommendations.

This review aimed to address this knowledge gap and has laid the foundations to achieve this. To our knowledge, this is the first review to explore the management of HMB in IBD. This review has highlighted the positive impact that readily available treatments can offer individuals experiencing HMB because of IBD, including both symptomatic relief and an improved QoL. Based on the data generated, guidance should be put into place whereby the Royal College of Obstetricians and Gynaecologists can act accordingly when encountering a patient with an IBD suffering from HMB, to manage the symptoms, improve patient outcomes and improve overall QoL. Ultimately, prior to considering any treatment, it is pertinent to address patients’ concerns about menorrhagia and bleeding disorders. Empowering patients so that they can actively manage their symptoms and find ways to tackle any unexplained symptoms helps and the World Federation of Hemophilia (31) and UKHCDO (9) are a source of guidance on this for patients and doctors.

As forementioned, FIGO note a non-structural cause of HMB to be IBD. However, in the NICE guidelines (1), the cause of HMB is not discussed; it is only mentioned to investigate for coagulation disorders if someone presents with HMB to rule it in or out as a cause. There is no guidance on how to treat and manage someone with an IBD who presents with HMB leaving a knowledge gap that can severely impact clinical practice and thus lead to negative or unwanted patient outcomes. In addition to this, the Royal College of Obstetricians and Gynaecologists (32) does not address the management of HMB in women with IBD. Despite this, the Haemophilia Society recognises that HMB is a common debilitating symptom of someone with a coagulation disorder and acknowledges that this can severely impact QoL. The Haemophilia Society set up a ‘Talking Red’ campaign (4) urging people to come together and talk about women’s bleeding to allow them to receive the treatment they require.

One of the studies (28) highlighted that, despite increased awareness of bleeding disorders in women with VWD over the past few decades, postpartum haemorrhage (PPH) remains a significant concern. Interestingly, improvements in care and updated guidelines have not reduced the incidence of PPH in these women. In cases where women received prophylactic treatment, the cause of PPH remains unclear. This issue ties into a bigger conversation about how women’s health, particularly menstruation and reproductive health, is often overlooked in mainstream discussions about mental health and productivity.

Therefore, our research has found that it is essential for future guidelines in this field to integrate menstrual management into workplace and school health programmes, ensuring that LNG-IUS is recognised within public health policies as a key tool for improving women’s overall well-being. It is crucial to balance effective treatment against side effects, to achieve a meaningful amelioration in the well-being of patients. It would also be interesting for studies to further explore the long-term mental health benefits, particularly in populations at risk for menstrual-related depression and anxiety.

In addition to expanding research, a concerted effort must be made to enhance public and professional awareness of menstrual health. Education on menstrual disorders, including inherited bleeding conditions, should be integrated into medical curricula, and continuing professional development programmes to ensure healthcare providers are better equipped to diagnose and manage these conditions. Furthermore, menstrual health education should be made more accessible to women of all ages, particularly in schools, workplaces, and community settings. Even public health campaigns can play a crucial role by addressing the stigma around menstrual health, ensuring that these issues are no longer side-lined but given the attention they deserve. Hopefully, by improving awareness and understanding, we can empower women to seek early intervention and take an active role in managing their health.

### Limitations

Although comprehensive this review had limitations. Certain articles were excluded by virtue of the exclusion criteria; including the screening out of case reports, reviews, and abstracts from conferences. This may have limited the generalisability of our results.

The inclusion of patients with IBD in some studies was not clear prior to reading the full article was read and were excluded prior to this step (33). To overcome this, we performed a final search on treatments and IBD and HMB, as well as a backward snowballing step, and found 35 articles that had been excluded during our original searches which were then fully assessed.

We started our literature search from 1^st^ January 2000 ensuring we encompassed almost 24 years of clinical experience of treating HMB in IBD patients. We could have searched further back in time but queried the relevance of such articles to current practices. We excluded studies involving paediatric populations (36, 37), as their physiological parameters differ significantly from adults, which could have introduced additional confounding factors. However menstruation often starts in childhood and would be first treated then. Indeed, some articles did not define how many patients were in each age group except that they included patients of <21 years of age.

While platelet function disorders can be both inherited and acquired, certain studies grouped all platelet function disorders together. This may have introduced variability in the included populations, potentially impacting the generalisability of our findings.

Additionally, although our MeSH terms were very comprehensive, there may be some obscurity on the definition of ‘abnormal uterine bleeding’ as our primary focus was HMB rather than abnormal bleeding. This could have restricted the results extracted from articles on clinical practice as the treatment strategies for AUB can differ depending on the underlying aetiology.

Another limitation was the lack of detailed patient outcome data in many studies, limiting our ability to extract outcome data from articles. Without access to raw data, we were unable to analyse all potential outcomes comprehensively, leading to gaps in our quantitative synthesis. In addition, the limited number of available studies and the scarcity of quantitative data constrained our ability to conduct statistical comparisons.

Lastly, we excluded certain articles (34, 35) because they were reviews rather than primary research studies. Our focus was on data from original experimental or clinical research to ensure the validity of any meta-analysis.

Despite these limitations, our study provides valuable insights into the treatment modalities for adolescents presenting with HMB. Future research with standardised definitions, detailed patient outcome reporting, and access to raw data would enhance the robustness of systematic reviews in this field.

### Recommendations

Addressing the knowledge gaps in the management of HMB, particularly in individuals with IBD, is essential for improving physical, mental and social well-being. Whilst there are recommendations for people with HMB, guidance for those with IBD remains limited. Below, we outline five key recommendations, informed via our research with proposed strategies for implementation in the clinical setting.

#### 1. LNG-IUS as first-line therapy

LNG-IUS is a highly effective treatment for HMB and should be considered as first-line treatment for individuals with IBD, given its ability to significantly reduce menstrual blood loss while providing long-term symptom control. While every patient should be assessed individually based on their symptoms to determine their suitability for LNG-IUS, there is growing evidence that introducing this therapy earlier in the treatment pathway, rather than as a last resort after other therapies have failed, leads to better symptom control and improved quality of life. Delaying its use until symptoms have significantly worsened may result in unnecessary prolonged distress for patients.

To implement this, gynaecologists should proactively discuss the LNG-IUS as an early treatment option, with training programmes emphasising shared decision-making to ensure patients understand the benefits and potential side effects of early LNG-IUS insertion. Additionally, improved access to specialist consultations in primary care settings would facilitate timely referrals and earlier intervention.

#### 2. The use of combination therapy

For patients in whom LNG-IUS is not suitable or insufficient, combination therapy with TA, desmopressin, or other agents should be explored. However, clinicians must remain cautious of venous thromboembolism (VTE) risks associated with certain treatments.

To implement this, a clear, evidence-based treatment algorithm should be developed, outlining when and how combination therapies should be used. It is pertinent to also incorporate risk stratification protocols into the clinical guidelines to ensure a considered balance between efficacy with safety.

#### 3. Strengthening counselling and communication between specialists

Managing HMB in individuals with IBD requires more than just prescribing the right treatment. Open, effective communication between specialists and a patient-centred approach to care is equally vital. Too often, patients find themselves caught between different specialties, with gynaecologists focused on managing menstrual symptoms and haematologists prioritising bleeding risks. A lack of co-ordination can lead to delays in treatment, conflicting advice, or even overlooked concerns that significantly impact a patient’s well-being.

To provide truly individualised care, discussions about treatment options should happen early and counselling should be incorporated to ensure patients feel heard, understood and involved in their own care. To successfully implement this, a centralised communication system can be developed which would help streamline care and ensure specialists are continually updated about a patient’s health. This could be further supported by clear national guidelines that encourage collaboration between gynaecologists, haematologists and GPs. More practically, simple steps like ensuring all patients have a named point of contact within each specialty and access to written treatment plans could make a tremendous difference to their confidence in managing their condition.

#### 4. Raising awareness through public and patient education

Education is key. Due to the lack of sufficient public awareness, many individuals with HMB delay seeking medical advice, assuming their symptoms are ‘just normal’ or something they must endure. This is even more pronounced for those with IBD, where a delayed diagnosis could potentially have serious long-term consequences. To combat this, a multifaceted approach needs to be designed.

As part of this approach, it will also be valuable to assess patients’ treatment compliance. As highlighted in the study by Dietricht et al. (17), iPods or similar technologies could be implemented to educate patients about their condition. Additionally, these devices can facilitate real-time data collection, enabling specialists to monitor patients’ health more effectively and promptly identify any untoward activity.

Moreover, clinics and hospitals could introduce more patient-friendly materials such as leaflets, digital resources and visual guides, targeted at those experiencing symptoms of HMB. Social media and community outreach programmes could also play a contributory role, particularly in reaching younger individuals who may not otherwise seek help. Collaboration with schools and primary care settings could further increase awareness, ensuring that conversations about menstrual health are normalised rather than stigmatised. It is indispensable to acknowledge that while these initiatives are crucial, we currently do not have concrete evidence proving that awareness campaigns directly lead to better outcomes. Nevertheless, equipping people with the knowledge to recognise their symptoms and seek timely care has the potential to make a profound difference, both in improving individual outcomes as well as reducing the burden of delayed diagnoses.

#### 5. Need for further research and discussion

Despite existing treatments, gaps remain in understanding the broader implications of HMB management in individuals with IBD. More research is needed on long-term treatment outcomes, side effects, the mental health impact of different therapies, and the logistical challenges of implementing new guidelines in the clinical settings. Future studies should prioritise patient-centred research, including longitudinal studies assessing treatment compliance, satisfaction and QoL outcomes. Research should also continuously explore innovative therapies and non-hormonal alternatives to provide more tailored treatment options.

Overall, there is a critical need for more focused research to optimise treatment options, promote early diagnosis, and improve overall management strategies.

HMB is both a symptom and condition within itself that should be better understood in theory through regular review so that its management can be better be explored in clinical practice. Based on the data extracted, guidance should be put into place whereby primary carers follow NICE guidelines (1) when encountering a patient with an IBD suffering from HMB. Our recommendations are based on current guidelines used by gynaecologists which should be shared with haematologists treating patients with IBDs.

## ACKNOWLEDGEMENTS

This work was funded in part through an INSPIRE grant awarded to the Hull York Medical School by the Academy of Medical Sciences through the Wellcome Trust [Ref: IR5\1018].

## Conflicts of Interest

The authors declare no conflicts of interest.

## Ethics

To ensure we met recognised standards of ethical considerations only articles that had followed their own countries ethical standards were included in this research.

## Authors contributions

Conception and design: J.T., D.A. and B.G.; resources: P.J.S. and H.S.; methodology and validation: M.G., K.F. and D.F.; analysis and interpretation of results, R.M., V.D., M.G., K.F. and B.G.; writing - original draft preparation: M.G., K.F., R.M. V.D. and B.G.; writing - review and editing: all authors; supervision: J.T, D.A. and B.G. All authors have read and agreed to the published version of the manuscript.

## Data Availability

The data generated and/or analysed during the current study are available as Supplementary Material at DOI: **10.5281/zenodo.15061468**.

## Supplementary Information

Supplementary Information I Protocol (Word document)

Supplementary Information II SLR (excel sheets)

Supplementary Information III Backward snowballing

Supplementary Information IV QA NOS

Supplementary Information V (A) Data Extraction (Excel); (B) Thematic analysis quotes and (C) Thematic Analysis Table

## Notes

### Competing Interest Statement

The authors have declared no competing interest.

### Author Declarations

The study used ONLY openly available human data that were originally located in articles citated in this article.

